# Knowledge and practice of preventive measures against COVID-19 infection among pregnant women in a low-resource African setting

**DOI:** 10.1101/2020.04.15.20066894

**Authors:** Johnbosco Ifunanya Nwafor, Joseph Kenechi Aniukwu, Bonaventure Okechukwu Anozie, Arinze Chidiebere Ikeotuonye

**Author notes:** ***Corresponding author:*** Johnbosco Ifunanya Nwafor, ***E-mail:***.

## Abstract

**Background:** Coronavirus disease pandemic has resulted in death of thousands of people across several countries. Several preventive measures have been recommended to halt the spread of the disease and its associated mortality. However, the level knowledge and practice of these preventive measures against COVID-19 infection among pregnant women, which constitute vulnerable groups, are yet to be evaluated.

**Aim:** To determine the knowledge and practice of preventive measures against COVID-19 infection among pregnant women in Abakaliki.

**Materials and Methods:** This was a self-administered questionnaire-based cross-sectional study conducted from February 1, 2020 to March 31, 2020 among 284 antenatal clinic attendees at Alex Ekwueme Federal University Teaching Hospital, Abakaliki, Ebonyi State. A pretested and validated questionnaire was used to collect the data. Data analysis was done using SPSS version 22.

**Results:** Of 284 participants, 60.9% (n=173) had adequate knowledge of the preventive measures against COVID-19 infection. However, the overall practice of these preventive measures among the participants were poor as 69.7% of the participants were not practicing the preventive measures against the coronavirus. The determinants of poor practice of the preventive measures among the participants were being in age group 31-40 years (AOR=2.04, 95%CI: 1.26 - 5.37, *p*=0.022), married (AOR=2.99, 95%CI: 1.40 - 6.33, *p*=0.035) grandmultiparous (AOR=3.11, 95%CI: 1.32 - 6.56, *p*=0.021), residing in rural area (AOR=2.08, 95%CI: 1.32 - 4.05, *p*=0.031), and having no formal education (AOR=6.73, 95%CI: 2.66 - 18.34, *p=*0.002).

**Conclusion:** The study showed that most of the participants had adequate knowledge of preventive measures against COVID-19 infection but the practice of these preventive measures were poor among the participants.

## Introduction

Emerging infections have been shown to have adverse impact on pregnant women and their fetuses as shown by recent pandemic caused by 2009 pandemic H1N1 influenza virus and the severe fetal effects of Zika virus.^1^

In December, 2019, a series of pneumonia cases of unknown cause emerged in Wuhan (Hubei, China), with clinical presentations resembling viral pneumonia. Deep sequencing analysis from lower respiratory tract samples indicated a novel coronavirus that was later named severe acute respiratory syndrome coronavirus 2 (SARS-CoV-2).^2^ The infection has spread to over 110 countries including Nigeria prompting World health organization to declare it a pandemic on March 11, 2020.^3^ Globally, as of April 15, 2020, there have been 1,914,916 confirmed cases of COVID-19, including 123,010 deaths, reported by WHO.^3^ Nigeria Centre for Disease Control announced the first confirmed case of coronavirus disease in Nigeria on February 27, 2020 and since then many confirmed cases have been reported in many States across the country.^4^

Early efforts have focused on describing the clinical characteristics and outcomes of Covid-19 in the general population.^5^ However, the disease appears to be particularly impactful in special populations that include those older than 65 years of age.^6^ Pregnant women are also considered to be a special population group because of the unique ‘immune suppression’ caused by pregnancy.^7^ The immunologic and physiologic changes of pregnancy might make pregnant women at higher risk of severe illness or mortality with Covid-19, compared with the general public.^6,7^ However, there is little information on Covid-19 infection during pregnancy.^7-9^ There are limited case series reporting the impact on women affected by coronaviruses (CoV) during pregnancy.^1,2,6^ In women affected by other coronavirus infections such as Middle East Respiratory Syndrome (MERS-CoV) and Severe Acute Respiratory Syndrome (SARS-CoV), the case fatality rate appeared higher in women affected in pregnancy compared with non-pregnant women. ^10^

To curtail the continued spread of the coronavirus disease and its associated mortality, World Health Organisation has recommended series of preventive measures including regular hand washing with water and soap, social distancing, covering hand and mouth while coughing and avoiding touching eyes, nose and mouth.^11^ In Nigeria, this preventive measures have been adopted to prevent further spread of the virus in the country. The government of Nigeria has also engaged in media campaigns to disseminate information on these preventive measures to the general public. However, the level of knowledge and practice of these preventive measures against COVID-19 infection among pregnant women, which constitute vulnerable groups, are yet to be evaluated. Therefore, this study aimed to determine the knowledge and practice of the preventive measures of coronavirus infection among pregnant women attending antenatal care at a tertiary hospital in Abakaliki, South-east, Nigeria.

## Materials and Methods

### Study design, period and area

This is a cross-sectional study that was conducted among pregnant women attending antenatal care at Alex Ekwueme Federal University Teaching Hospital, Abakaliki from February 1, 2020 to March 31, 2020. Ebonyi state is one of the five states in the South-east geopolitical zone of Nigeria.^12^ Abakaliki is a semi-urban area and the capital of Ebonyi State, Nigeria. The population comprises mainly of subsistence farmers and petty traders. Christianity forms their major religion and the inhabitants are predominantly Igbo speaking.

### Study population

All consenting eligible pregnant women who attended antenatal care during the study period.

### Study criteria

All women who gave informed consent to participate in the study were included in the study. Participants who had a verbal communication problem and complete loss of hearing were excluded.

### Sample size determination

The sample size was calculated by taking variability of proportion of 18% from the average of 450 pregnant women who attend antenatal clinic per month in the hospital.^13^ Using Open Epi software package for the determination of sample size and design effect of 1.5 at error margin of 5%, the final minimum sample size was calculated as 227. After considering 10% non-response rate for any unpredictable events, the final required sample size was 249.7. However, 284 women were recruited for the study.

### Sampling technique

The study participants were selected by using simple random sampling method and the first participant was selected by using lottery method.

### Study variables

The dependent (outcome) variables for this study were the level of knowledge and practice of preventive measures against COVID-19 infection. The independent variables were: age, parity, marital status, educational attainment, area of residence, occupation and husband’s educational level.

### Operational definition

#### Awareness

A participants were classified as being aware of coronavirus disease pandemic if a positive response (‘Yes’) is obtained to the question ‘have you ever heard of coronavirus disease pandemic?’

#### Adequate knowledge

Participants who scored ≥ 65% (score 8 - 12) on knowledge of coronavirus disease preventive measures questionnaire.

#### Inadequate knowledge

Women who scored < 60% (score up to 6) on knowledge of coronavirus disease preventive measures questionnaire.

#### Good practice

Women who scored 100% (score of 12) on the practice of coronavirus disease preventive measures questionnaire.

#### Poor practice

Women who scored < 100% (score below 12) on the practice of coronavirus disease preventive measures questionnaire.

### Data collection tools and procedures

A self-administered questionnaire on the knowledge and practice of coronavirus infection preventive measures was used for data collection. The questionnaire was developed following review of literatures on the WHO recommendations on the measures to prevent human-to-human transmission of COVID-19 infection.^11^ The study questionnaire was divided into four areas: 1)sociodemographic characteristics (ie, age, parity, marital status, area of residence, occupation, participant’s level of education, and husband’s level of education); 2)media exposure to access information about coronavirus infection pandemic; 3)knowledge about preventive measures against Coronavirus (COVID-19) infection (ie, ever heard about coronavirus infection pandemic, multiple variables regarding symptoms, what to do when the participants suspect to have develop symptoms and preventive measures of coronavirus infection); 4) Practice of preventive measures against coronavirus infection. The variables used to assess the knowledge and practice of preventive measures of coronavirus infection were 1) Frequent hand washing with soap and water or rubbing alcohol-based sanitizers on the hand, 2) Maintaining at least 1 meter distance from others, 3) Avoiding touching eyes, nose and mouth with hands, 4) Covering mouth and nose when coughing or sneezing, 5) Wearing face mask in public, 6) Staying indoor.

The questionnaire has 12-item scale (6-item for knowledge questions and 6-item for practice questions). The scoring system of women’s knowledge and practice of preventive measures was either 2 (for correct answer) or 0 (for incorrect answer). The minimum score was 0 whereas the maximum score was 12 each for both knowledge and practice component of the questionnaire.

The reliability of the questionnaire was checked by conducting a pretest among pregnant women in the antenatal clinic, by taking 5% of the sample size. From the pretest, understandability, clarity, and organization of the questionnaire were checked. From the reliability test of knowledge and practice questions, 0.898 Cronbach’s alpha value was found. The questionnaire was prepared in English language and then translated to Igbo (local language in Abakaliki) that was used for data collection and re-translated back to English to check its consistencies. The questionnaire was then refined accordingly for final use. Two trained house officers participated in data collection. The data collector were trained for one day on the techniques of data collections. The training also included the importance of disclosing the possible benefit and purpose of the study to the study participants before the start of data collection.

### Data quality control and analysis

The principal researcher checked completeness and consistency of questionnaires filled by the data collectors to ensure the quality of data. The collected data were entered and analyzed using SPSS version 22 (IBM Corp. Amork, New York, U.S.A). Proportions, rates and summary statistics such as mean, the standard deviation were calculated for most variables. Bivariate regression analysis was employed to test the strength of the association between maternal socio-demographic characteristics and knowledge and practice of preventive measures of coronavirus infection, and the results were expressed as crude odds ratio (COR) with the corresponding 95% confidence interval. The variables that showed significant association with knowledge and practice of preventive measures of coronavirus infection were included in multivariate logistic regression model to identify the independent predictors of poor knowledge and practice of preventive measures of coronavirus infection. The results of the multivariate regression analysis were expressed as adjusted odds ratio (AOR) at 95% confidence interval, with level of significance set at p < 0.05.

### Ethical considerations

This study was approved by the Research and Ethics Committee (REC) of the Alex Ekwueme Federal University Teaching Hospital, Abakaliki. Informed consent was taken from the study participants after informing the study subjects on study objectives, expected outcomes, and benefits associated with it. Confidentiality of responses was maintained throughout the study.

## Results

A total of 284 pregnant women participated in the study and all the study participants (100%) were aware of the COVID-19 infection global pandemic. The mean age of the study cohorts was 24.6 ± 6.3 years, ranged from 18 to 42 years. Majority of the participants were married (78.9%), multiparous (39.4%), farmers (34.9%) and reside in urban area (60.9%). Most of the participants (89.8%) and their husbands (94.4%) had some formal education.

The depth of knowledge of preventive measures against coronavirus infection among the participants is shown in Figure 1. Over half (60.9%) of the participants had adequate knowledge of the preventive measures, whereas 39.1% of the women were classified as having inadequate knowledge of the preventive measures against coronavirus infection.

**Figure 1:**
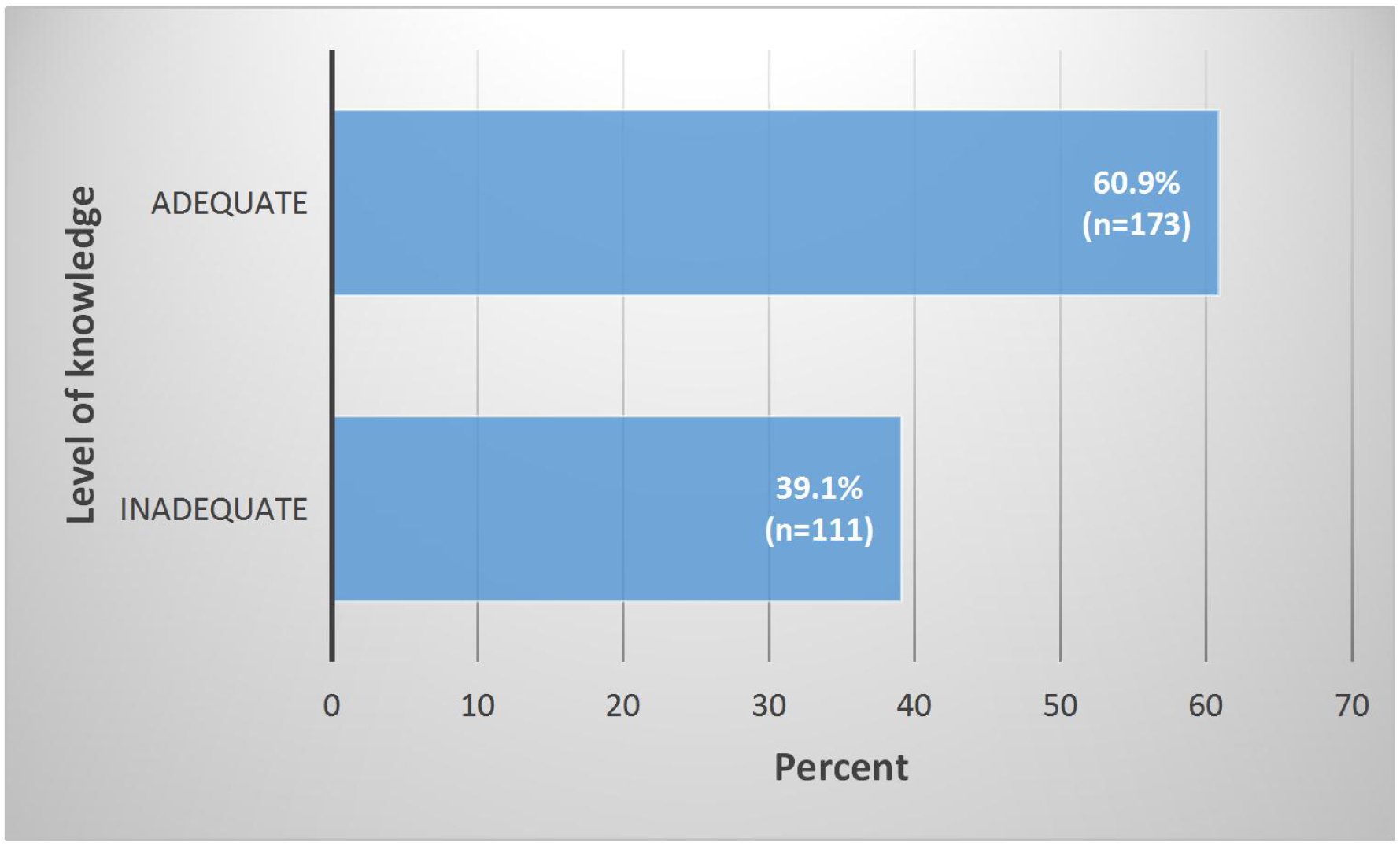
Level of knowledge of preventive measures against COVID-19 infection among the participants.

Figure 2 shows the number of the participants that adopted the preventive measures against coronavirus infection transmission. Most (69.7%) of the participants’ practices to prevent coronavirus infection transmission were within poor practice category. Only 30.3% of the participants’ practices to prevent human-to-human transmission of the virus were within good practice scores.

**Figure 2:**
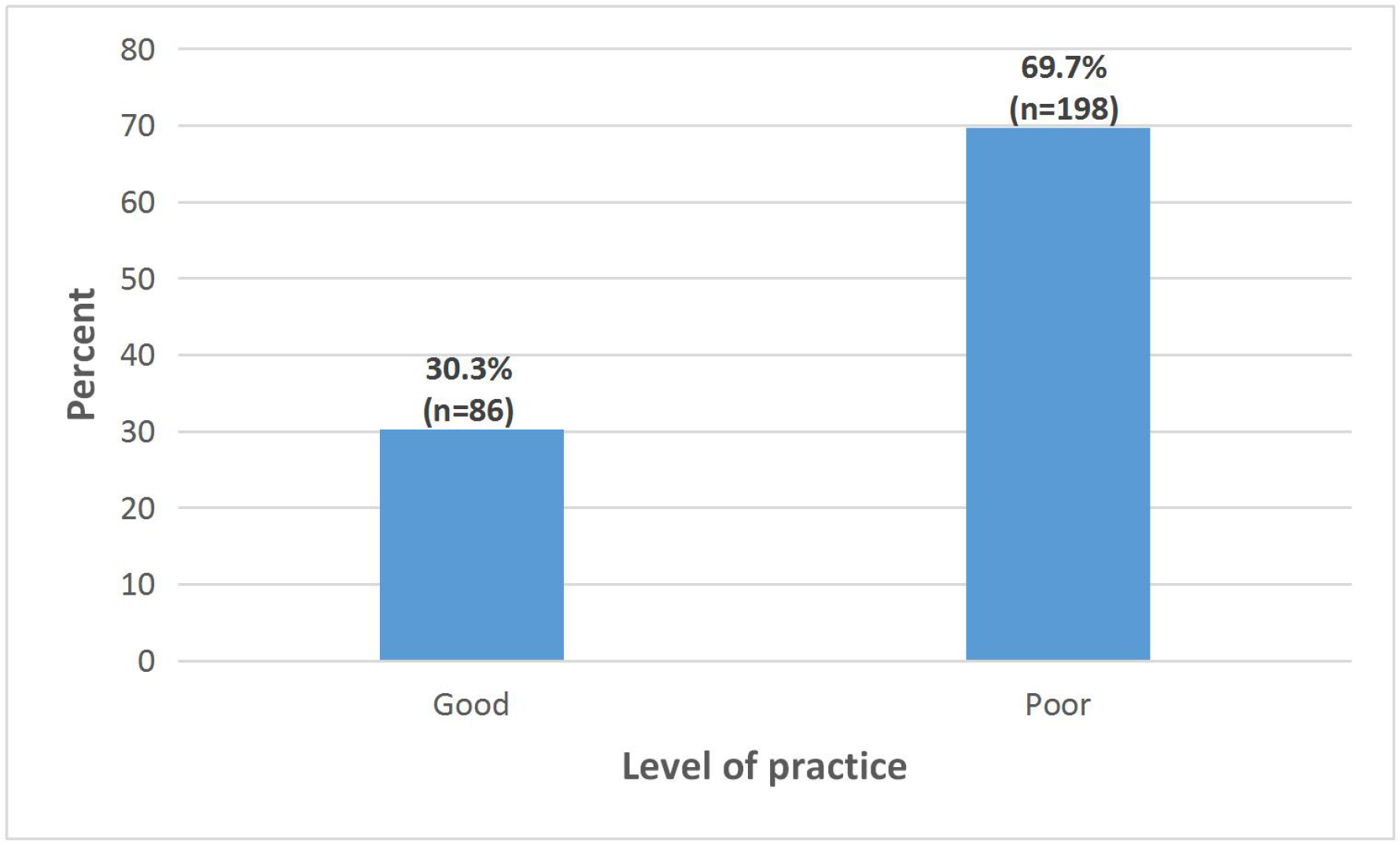
Level of practice of preventive measures against COVID-19 infection among the participants.

The sources of information on coronavirus infection pandemic for the study participants is shown in Table 2. The commonest source of information for the participants was the television (82.7%), followed by friends and relatives (78.5%). Health workers and internet accounted for 39.8% and 44.7% of sources of information for the participants respectively.

**Table 1:** Sociodemographic characteristics of the study participants (N = 284)

| Variable | Frequency (%) |
| --- | --- |
| Age (years) |  |
| 18 - 22 | 56 (19.7) |
| 23 - 27 | 109 (38.4) |
| 28 - 32 | 37 (13.0) |
| 33 - 37 | 45 (15.8) |
| 38 - 42 | 37 (13.1) |
| Mean age ± SD (years) | 24.6 ± 6.3 |
| 0 | 56 (19.7) |
| 1 | 59 (20.8) |
| 2-4 | 112 (39.4) |
| $\geq 5$ | 57 (20.6) |
| Mean parity ± SD | 2.1 ± 1.4 |
| Marital status |  |
| Married | 224 (78.9) |
| Single | 29 (10.2) |
| Divorced | 12 (4.2) |
| Widowed | 19 (6.7) |
| Educational attainment |  |
| No formal education | 29 (10.2) |
| Primary | 67 (23.6) |
| Secondary | 107 (37.7) |
| Tertiary | 81 (28.5) |
| Area of residence |  |
| Urban | 173 (60.9) |
| Rural | 111 (39.1) |
| Occupation |  |
| Farming | 99 (34.9) |
| Artisan | 34 (11.9) |
| Trading | 70 (24.6) |
| Civil servant | 56 (19.7) |
| Unemployed | 25 (8.9) |
| Husband's educational attainment |  |
| No formal education | 16 (5.6) |
| Primary | 100 (35.2) |
| Secondary | 99 (34.9) |
| Tertiary | 69 (24.3) |

**Table 2:** Sources of information on preventive measure against COVID-19 infection

| Source of information* | Frequency (%) |
| --- | --- |
| Health workers | 113 (39.8) |
| Friends and relatives | 223 (78.5) |
| Television | 235 (82.7) |
| Radio | 189 (66.5) |
| Internet | 127 (44.7) |
| *Multiple responses |  |

Participants’ responses of knowledge of coronavirus infection and its preventive measures are shown in Table 3. Cough (75.7%) and difficult breathing (82.4%) were most common symptoms known by participants. Just over half (53.9%) of the participants knew that fever is a symptom of coronavirus infection. Majority of the participants had inadequate knowledge of what to do when they suspect to have developed symptoms of coronavirus infection. Fifty-seven (20.1%) women said they would stay at home, 15.8% reported that they would wear facemask and 4.2% said they would inform their health care provider in advance before visit to hospital. Significant number of the study cohorts had adequate knowledge of preventive measures to prevent human-to-human transmission of coronavirus infection. The preventive measures known by participants were washing hands frequently with soap and water or rubbing hands with alcohol-based sanitizers (93.7%), maintaining at least 1 meter distance between yourself and others (87.7%), avoiding touching eyes, nose and mouth with hands (75%), covering mouth and nose when coughing or sneezing (97.5%), wearing facemask in public (98.6%) and staying indoor (74.3%).

**Table 3:** Participants’ responses of knowledge towards COVID-19 infection and its prevention measures

| Variables | Yes<br>n(%) | No<br>n(%) |
| --- | --- | --- |
| **Are you aware of ongoing COVID-19 infection pandemic?** | 284 (100) | 0 (0) |
| **What are the symptoms of COVID-19 infection?\*** |  |  |
| Fever | 153 (53.9) | 131 (46.1) |
| Headache | 89 (31.3) | 195 (68.7) |
| Runny nose | 95 (33.5) | 189 (66.5) |
| Cough | 215 (75.7) | 69 (24.3) |
| Difficulty breathing | 234 (82.4) | 50 (17.6) |
| **What will you do when you have the above symptoms?\*** |  |  |
| Stay at home | 57 (20.1) | 227 (79.9) |
| Wear face mask | 45 (15.8) | 239 (84.2) |
| Inform your health care provider in advance before visit to hospital | 12 (4.2) | 272 (95.8) |
| **What are the preventive measure of COVID-19 infection?\*** |  |  |
| Washing hands frequently with soap and water or rubbing hands with alcohol-based sanitizers | 266 (93.7) | 18 (6.3) |
| Maintaining at least 1 meter distance between yourself and others | 249 (87.7) | 31 (12.3) |
| Avoid touching eyes, nose and mouth with hands | 213 (75.0) | 71 (25.0) |
| Covering mouth and nose when coughing or sneezing | 277 (97.5) | 7 (2.5) |
| Wearing facemask in public | 280 (98.6) | 4 (1.4) |
| Staying indoor | 211 (74.3) | 73 (25.7) |

\*Multiple response

Table 4 shows the participants’ responses toward practice of preventive measures against COVID-19 infection. Seventy-six (26.8%) participants practised frequent hand washing with soap and water while 20.4% practised social distancing of at least 1 meter between them and others. Avoidance of touching eyes, nose and mouth with hands was practised by 21.5% of the study cohort whereas 32.7% of the participants used facemask in public.

**Table 4:** Participants’ responses toward practice of preventive measures of COVID-19 infection

| Variable | Yes<br>n(%) | No<br>n(%) |
| --- | --- | --- |
| **What measures did you adopt to prevent COVID-19 infection?\*** |  |  |
| Washing hands frequently with soap and water or alcohol-based sanitizers | 76 (26.8) | 208 (73.2) |
| Maintaining at least 1 meter distance between yourself and others | 58 (20.4) | 226 (79.6) |
| Avoid touching eyes, nose and mouth with hands | 61 (21.5) | 223 (78.5) |
| Wearing face mask in public | 93 (32.7) | 191 (67.3) |
| Staying indoors | 49 (17.3) | 235 (82.7) |
| *Multiple response |  |  |

The determinants of knowledge of preventive measures against coronavirus infection among the participants is shown in Table 5. Age, parity, level of educational attainment, area of residence and occupation were factors associated with inadequate knowledge of preventive measures against COVID-19 infection among the participants. Women who were > 40 years (AOR=0.19, 95%CI: 0.23 - 0.65, *p* <0.001) were less likely to have adequate knowledge when compared with younger age groups. On the other hand, women who were above para 5 were less likely to have inadequate knowledge of preventive measures against coronavirus infection (AOR=0.24, 95%CI: 0.21 - 0.87, *p*=0.002). Participants who had no formal education were 6 times more likely to have inadequate knowledge of preventive measures when compared with those with formal education (AOR=6.30, 95%CI: 2.55 - 6.91, *p=*0.004). Similarly, women residing in rural areas were 9 times more likely to have inadequate knowledge of preventive measures when compared with participants living in urban areas (AOR=9.11, 95%CI: 5.67 - 20.01, *p*<0.001). Also, women who were artisans were about thrice more likely to have inadequate knowledge of preventive measures (AOR=2.82, 95%CI: 0.02 - 0.77, *p=*0.021).

**Table 5:** Predictors of knowledge of preventive measures against COVID-19 infection among study participants

| Variable | Knowledge |  | COR(95% CI) | *p* value | AOR(95% CI) | *p* value |
| --- | --- | --- | --- | --- | --- | --- |
|  | Adequate | Inadequate |  |  |  |  |
|  | (n=173) | (n=111) |  |  |  |  |
| Age (years) |  |  |  |  |  |  |
| < 30 | 76 | 70 | 1.00 |  | 1.00 |  |
| 31 - 40 | 54 | 31 | 0.62 (0.36 - 1.08) | 0.091 |  |  |
| > 40 | 43 | 10 | 0.25 (0.12 - 0.54) | <0.001 | 0.19 (0.23 - 0.65) | <0.001 |
| Parity |  |  |  |  |  |  |
| 0 | 32 | 24 | 1.00 |  | 1.00 |  |
| 1 | 49 | 10 | 0.27 (0.11 - 0.64) | 0.003 | 0.89 (0.22 - 3.45) | 0.491 |
| 2-4 | 41 | 71 | 2.31 (1.20 - 4.44) | 0.012 | 1.01 (0.40 - 5.88) | 0.425 |
| ≥ 5 | 51 | 6 | 0.16 (0.05 - 0.43) | <0.001 | 0.24 (0.21 - 0.87) | 0.002 |
| Marital status |  |  |  |  |  |  |
| Single | 12 | 17 | 1.00 |  | 1.00 |  |
| Married | 140 | 84 | 0.42 (0.19 - 0.93) | 0.032 | 0.76 (0.56 - 1.72) | 0.151 |
| Divorced | 6 | 6 | 0.71 (0.18 - 2.73) | 0.614 |  |  |
| Widowed | 15 | 4 | 0.19 (0.05 - 0.71) | 0.014 | 0.39 (0.63 - 1.85) | 0.413 |
| Educational attainment |  |  |  |  |  |  |
| No formal education | 4 | 25 | 50.6 (44.07 - 67.39) | <0.001 | 6.30 (2.55 - 6.91) | 0.004 |
| Primary | 32 | 35 | 1.45 (0.43 - 3.45) | 0.652 |  |  |
| Secondary | 57 | 50 | 2.12 (0.52 - 3.22) | 0.593 |  |  |
| Tertiary | 80 | 1 | 1.00 |  | 1.00 |  |
| Area of residence |  |  |  |  |  |  |
| Urban | 141 | 32 | 1.00 |  | 1.00 |  |
| Rural | 32 | 79 | 10.88 (6.20 - 19.08) | <0.001 | 9.11 (5.67 - 20.01) | <0.001 |
| Occupation |  |  |  |  |  |  |
| Farming | 49 | 50 | 0.68 (0.29 - 1.66) | 0.397 |  |  |
| Artisan | 27 | 7 | 2.17 (0.05 - 0.54) | 0.003 | 2.82 (0.02 - 0.77) | 0.021 |
| Trading | 36 | 34 | 0.63 (0.25 - 1.59) | 0.328 |  |  |
| Civil servant | 51 | 5 | 0.57 (0.32 - 0.52) | 0.01 | 1.33 (1.11 - 5.40) | 0.231 |
| Unemployed | 10 | 15 | 1.00 |  | 1.00 |  |
| Husband's educational level |  |  |  |  |  |  |
| No formal education | 5 | 11 | 1.03 (1.55 - 16.28) | 0.047 | 1.11 (1.69 - 18.20) | 0.055 |
| Primary | 43 | 57 | 3.03 (1.59 - 5.79) | 0.041 | 1.95 (0.46 - 3.82) | 0.063 |
| Secondary | 77 | 22 | 0.65 (0.32 - 1.31) | 0.232 |  |  |
| Tertiary | 48 | 21 | 1.00 |  | 1.00 |  |

Factors associated with poor practice of preventive measures against COVID-19 infection are shown in Table 6. The odds of poor practice of preventive measures was 2 times higher in women between the age of 31 and 40 years when compared with those younger than 30 years (AOR=2.04, 95%CI: 1.26 - 5.37, *p*=0.022). Grandmultiprous (para ≥ 5) (AOR=3.11, 95%CI: 1.32 - 6.56, *p*=0.021) participants were 3 times respectively more likely not to practice preventive measures against coronavirus infections when compared with nulliparous women. When compared with women who were single, the odds of poor practice of preventive measures were almost 3 times higher among married participants (AOR=2.99, 95%CI: 1.40 - 6.33, *p*=0.035). The odds of poor practice of preventive measures of coronavirus infection decrease as educational attainment increases. Participants who had no formal education (AOR=6.73, 95%CI: 2.66 - 18.34, *p*=0.002) were approximately 7 times more likely to have poor practice scores of preventive measures when compared to those with tertiary education. Similarly, when compared with participants with tertiary education, those who had primary educational were nearly 6 times more likely to have poor practice scores (AOR=5.95, 95%CI: 3.02 - 14.39, *p*<0.001). Area of residence was a significant determinants in the practice of preventive measures. Women residing in rural areas had the odds of poor practice of 2 times higher (AOR=2.08, 95%CI: 1.32 - 4.05, *p=*0.031) when compared with those living in urban areas. Participants who were farmers (AOR=10.05, 95%CI: 3.89 - 28.03, *p*<0.001), artisans (AOR=2.99, 95%CI: 1.30 - 10.93, *p*=0.03) and traders (AOR=5.87, 95%CI: 2.32 - 12.64, *p*=0.002) were unlikely to adopt preventive measures against coronavirus infection.

**Table 6:** Predictors of poor practice of preventive measures against COVID-19 infection among study participants

| Variable | Practice |  | COR(95% CI) | *p* value | AOR(95% CI) | *p* value |
| --- | --- | --- | --- | --- | --- | --- |
|  | Good | Poor |  |  |  |  |
|  | (n=86) | (n=198) |  |  |  |  |
| Age (years) |  |  |  |  |  |  |
| < 30 | 43 | 83 | 1.00 |  | 1.00 |  |
| 31 - 40 | 27 | 94 | 1.80 (1.03 - 3.17) | 0.041 | 2.04 (1.26 - 5.37) | 0.022 |
| > 40 | 16 | 21 | 0.68 (0.32 - 1.44) | 0.312 |  |  |
| Parity |  |  |  |  |  |  |
| 0 | 29 | 27 | 1.00 |  | 1.00 |  |
| 1 | 21 | 38 | 1.94 (0.92 - 4.10) | 0.081 |  |  |
| 2-4 | 19 | 93 | 5.26 (2.56 - 10.79) | <0.001 | 4.01 (0.55 - 6.88) | 0.601 |
| ≥ 5 | 17 | 40 | 2.53 (1.17 - 5.47) | 0.019 | 3.11(1.32 - 6.56) | 0.021 |
| Marital status |  |  |  |  |  |  |
| Single | 13 | 16 | 1.00 |  | 1.00 |  |
| Married | 56 | 168 | 2.44 (1.10 - 5.38) | 0.027 | 2.99 (1.40 - 6.33) | 0.035 |
| Divorced | 5 | 7 | 1.14 (0.29 - 4.43) | 0.853 |  |  |
| Widowed | 12 | 7 | 0.47 (0.14 - 1.55) | 0.217 |  |  |
| Educational attainment |  |  |  |  |  |  |
| No formal education | 5 | 24 | 7.35 (2.54 - 21.25) | <0.001 | 6.73 (2.66 - 18.34) | 0.002 |
| Primary | 13 | 54 | 6.36 (2.99 - 13.49) | < 0.001 | 5.95 (3.02 - 14.39) | <0.001 |
| Secondary | 19 | 88 | 7.09 (3.64 - 13.81) | < 0.001 | 3.68 (0.87 - 6.44) | 0.051 |
| Tertiary | 49 | 32 | 1.00 |  | 1.00 |  |
| Area of residence |  |  |  |  |  |  |
| Urban | 61 | 112 | 1.00 |  | 1.00 |  |
| Rural | 25 | 86 | 1.86 (1.08 - 3.19) | 0.026 | 2.08 (1.32 - 4.05) | 0.031 |
| Occupation |  |  |  |  |  |  |
| Farming | 15 | 84 | 9.96 (3.72 - 26.64) | <0.001 | 10.05 (3.89 - 28.03) | <0.001 |
| Artisan | 11 | 23 | 3.71 (1.25 - 11.03) | 0.018 | 2.99 (1.30 - 10.93) | 0.03 |
| Trading | 17 | 53 | 5.54 (2.07 - 14.81) | <0.001 | 5.87 (2.32 - 12.64) | 0.002 |
| Civil servant | 27 | 29 | 1.91 (0.72 - 5.04) | 0.191 |  |  |
| Unemployed | 16 | 9 | 1.00 |  | 1.00 |  |
| Husband's educational level |  |  |  |  |  |  |
| No formal education | 6 | 10 | 2.93 (0.95 - 9.03) | 0.061 |  |  |
| Primary | 20 | 80 | 3.04 (1.52 - 4.09) | 0.036 | 1.89 (0.37 - 2.55) | 0.059 |
| Secondary | 16 | 83 | 2.13 (1.42 - 5.87) | 0.05 |  |  |
| Tertiary | 44 | 25 | 1.00 |  | 1.00 |  |

## Discussion

This study provides an insight on the level of knowledge and practice of preventive measures against person-to-person transmission of coronavirus infection among pregnant mothers in Abakaliki at the time of the COVID-19 infection outbreak in Nigeria. This study is unique because it is the first study to describe the knowledge and adoption of preventive practices to protect pregnant mothers from coronavirus disease and its associated morbidity and mortality.

In this study, the level of knowledge about preventive measures against coronavirus disease among the participants was high. This is probably because since the onset of the first confirmed case of the disease, Nigerian government have embarked on aggressive media campaign to educate the populace on the preventive measures to curtail person-to-person transmission of the disease. Therefore, it is not surprising that television and friends and relatives were the sources of information on the preventive measures for the majority of the study participants.

Although, most of the study participants had adequate knowledge about the preventive measures to halt the spread of the disease, the level of practice of these preventive measures remained poor. This could be attributed to sociodemographic characteristics of the population in sub-Saharan African countries like Nigeria. High parity, rural residence, low educational attainment and occupations requiring physical contacts were the factors associated with poor practice of preventive measures against the disease. These factors increase pregnant women’s risk of exposure and contracting coronavirus disease as it continued to spread in Nigeria.

Because COVID-19 is an emerging infectious disease, the optimal treatment for affected individuals has not yet been established.^1,2,6^ Currently, the effect of coronavirus infection in pregnancy is not well described.^14^ It is important that vulnerable populations such as pregnant women be protected from coronavirus infection. This is particularly of utmost importance especially in sub-Saharan African countries like Nigeria where the health infrastructure is weak and could not handle coronavirus in massive proportion as seen in developed countries who are struggling to contain the disease with its associated mortality in the setting of advanced health systems.

About 295, 000 women died during and following pregnancy and childbirth in 2017 and 94% of these deaths occurred in low-resource settings.^15^ Nigeria contributes nearly 20% of global maternal deaths.^16^ Nigeria’s maternal mortality ratio is over 800 maternal deaths per 100, 000 live births and most of these deaths are preventable.^16^ Therefore, poor practices of preventive measures against COVID-19 infection among pregnant women would put these women at high risk of infection which could worsen our maternal morbidity and mortality profile during this coronavirus pandemic. Although, case reports from advanced countries about the outcome of coronavirus disease in pregnancy appear to be good^8^ and and these outcomes were achieved with intensive, active management that might be absent in most developing countries due to poor healthcare system prevalent in resource-constrained settings. Therefore, pregnant women require special attention in relation to prevention, diagnosis, and management.

In order to effectively contain the spread of the coronavirus infection to Nigerian population and to protect vulnerable populations such as pregnant women, this current media campaign should be extended to rural areas where access to electronic media is limited. In addition, provision of economic palliative support to families who depend on daily income for survival would likely encourage women, who in some cases are the breadwinners in many households, to adopt preventive measures to halt the spread of this virus in Nigeria.

## Strengths and limitation

The strength of this study is that it is the first study to evaluate the knowledge, practice and sociodemographic variables associated with poor knowledge and practice of preventive measures against coronavirus infection among pregnant mothers in a low-resource setting. These associated factors of poor practice were determined using bivariate and multivariate logistic regression analysis. These statistical methods are appropriate and efficient method for determining associations between dependent and independent variables. Despite these strengths, this study has a limitation. It is a single centre study which limits the generalization of the study findings to the study area. A multicentre study would have been ideal.

## Conclusion

In conclusion, this study showed that most of the participants had adequate knowledge of preventive measures against COVID-19 infection. However, the practice of these preventive measures were poor among the participants. Multiparity, rural residence, low level of educational attainment and occupations (such as farming, trading and artisan) were factors significantly associated with poor practice of the preventive measures against coronavirus -19 infection among pregnant women.

## Data Availability

The data is available from the corresponding author on request

## Acknowledgement

None

